# Unusual trend of respiratory syncytial virus burden of disease observed in primary care diagnosis of children under 5 years old in Catalonia during the COVID-19 pandemic

**DOI:** 10.1101/2021.01.27.21250063

**Authors:** Ermengol Coma, Jorgina Vila, Leonardo Méndez-Boo, Andrés Antón, Núria Mora, Francesc Fina, Mireia Fàbregas, Manuel Medina

## Abstract

We observed an unusual pattern of respiratory syncytial virus (RSV) in children under 5 years in Catalonia (Spain). We observed a nearly absence during winter months and a subsequent surge late spring. Primary care electronic health records combined with hospital RSV laboratory confirmations could be a useful surveillance system to monitorize trends of respiratory pathogens.

## Introduction

Respiratory syncytial virus (RSV) is the main etiological agent of acute bronchiolitis, the most common lower respiratory tract infection in infancy and the leading cause of morbidity in infants less than one year old [1]. In Catalonia (Spain), the RSV epidemic occurs every year between November and March, and usually peaks during late December, before the influenza epidemic [2].

During the 2020 Southern hemisphere winter, Yeoh et al. reported a 98% reduction of RSV in Australia coinciding with the COVID-19 pandemic [3]. Later on, a subsequent spring and summer surge in RSV activity was described from late September in some Australian regions [4]. The unusual evolution was related to the impact of non-pharmaceutical interventions such as physical distancing measures and respiratory hygiene practices during the COVID-19 pandemic according to the authors’ conclusions. The winter wave in 2021 was also absent in different european countries[5] and there were some concerns about the possibility of a delayed RSV peak.

Some studies have suggested that clinical diagnoses from digital records could be useful for the RSV surveillance [6]. In Catalonia, most of the primary care practices use the same electronic health record system (EHR), covering > 6 million people (88% of the Catalan population). EHR data have been previously used in some analysis of respiratory pathogens trends during COVID-19 pandemic [7].

We aim to describe the current trend of annual RSV epidemic and the associated respiratory disease in children in Catalonia compared to the previous ten years in the context of the COVID-19 pandemic, using data from primary care EHR as a clinical surveillance system for RSV related disease and supported with data from hospital microbiology laboratory.

## Methods

Time series analysis of RSV clinical diagnoses in all primary care settings in Catalonia and RSV laboratory-confirmations performed in Hospital Universitari Vall d’Hebron (Barcelona, Catalonia, Spain), which is one of the largest hospital complexes in Spain. Daily counts of RSV-related disease were retrospectively extracted from the primary care electronic health records (EHR). The study period included 12 epidemiological seasons from 1 September 2009 (2009-2010 season) to 11 July 2021 (2020-2021 season).

We included all children younger than 5 years old with a clinical diagnosis of suspected RSV infection according to ICD-10 codes registered in the EHR: J21 (Acute bronchiolitis), J21.0 (Acute bronchiolitis due to RSV), J21.9 (Acute bronchiolitis, unspecified) and B97.4 (Respiratory syncytial virus as the cause of diseases classified elsewhere). Hospital laboratory-confirmations for RSV and other respiratory viruses in respiratory specimens from children <5 years old who were attended at Vall d’Hebron hospital were routinely performed by using direct immunofluorescence antigen detection (D3 Ultra 8™ DFA Respiratory Virus Screening & Identification Kit, Diagnostic HYBRIDS, USA) or real-time multiplex RT-PCR (Anyplex™ II RV16 Detection kit, Seegene, South Korea, during the 2014-2015 season; and Allplex™ Respiratory Panel, Seegene, South Korea, from 2015-2016 to 2017-2018). Additionally, the detection of RSV during epidemics with rapid tests was performed using an immunofluorescent assay (Sofia RSV FIA, Quidel, CA, USA) or a rapid molecular test (GeneXpert Flu or Flu/RSV XC, Cepheid, CA, USA). If the rapid test was negative, the study was extended to the routine diagnosis of other respiratory viruses, as described in Gimferrer et al [2].

## Statistical analysis

Daily counts of RSV cases were computed based on the frequency of cases recorded in the previous 7-day period to avoid weekly effects on recording practice.

We obtained the expected cases for the study period using a time series regression adjusted by seasonality. Dataset was divided into two sets: training set (from September 2009 to August 2019) and analysis set (from September 2019 to July 2021). We used the training set to adjust the model and we projected the estimated time series to our analysis set. We calculated a 95% confidence interval (CI) for each estimate. Reduction or excess of RSV was defined as the difference between the expected minus the observed for all periods in which the observed number was outside the 95% CI.

We performed the same time-series analysis with RSV laboratory-confirmation from Hospital Universitari Vall d’Hebron with data available between week 40/2013 and week 26/2021 (previous data were missing). Weekly counts of RSV cases were computed using the same method described above.

All analyses were performed in R v.3.5.1.

## Results

From 1 September 2009 to 11 July 2021, we observed 196,277 RSV related diseases of whom 184,383 (94%) were diagnosed in the population under two years of age and 42.2% were female. The average number of RSV cases for seasons 2009-2010 to 2018-2019 was 17,605; for season 2019-2020 was 13,540 but only 6,686 for the 2020-2021 season (until 11 July 2021). Numbers of new RSV-related disease cases by age groups are shown in Supplementary Table S1, while Supplementary Figure S1 presents the trends since 2009.

Figure 1 shows the observed and estimated number of 7-day new RSV-related diseases (with 95% CI) for the analysis set. We observed that RSV cases remain between the 95% CI during the 2019-2020 winter, with lower observed cases than expected in March, April and May 2020, after the state of alarm was setted in Spain on March 14th. However, during the 2020-2021 period, RSV cases were lower than expected throughout all winter months. Overall, the number of RSV-related disease diagnoses accounted for 11,499 fewer diagnoses resulting in a 86.1% reduction from expected (95% CI: 79.1% to 89.6%) from 7 October 2020 to 26 March 2021. Afterwards, we observed a resurgence of RSV-related disease from mid-April, resulting in 2,014 more cases until July 11 (end of the study). Although this increase accounted for a 279% excess for that period, its peak was roughly a half of those usually occurring in winter. At the time of writing, the current wave is already descending but still higher than the expected 95% upper CI so the percentage of excess will be higher.

**Figure 1.**
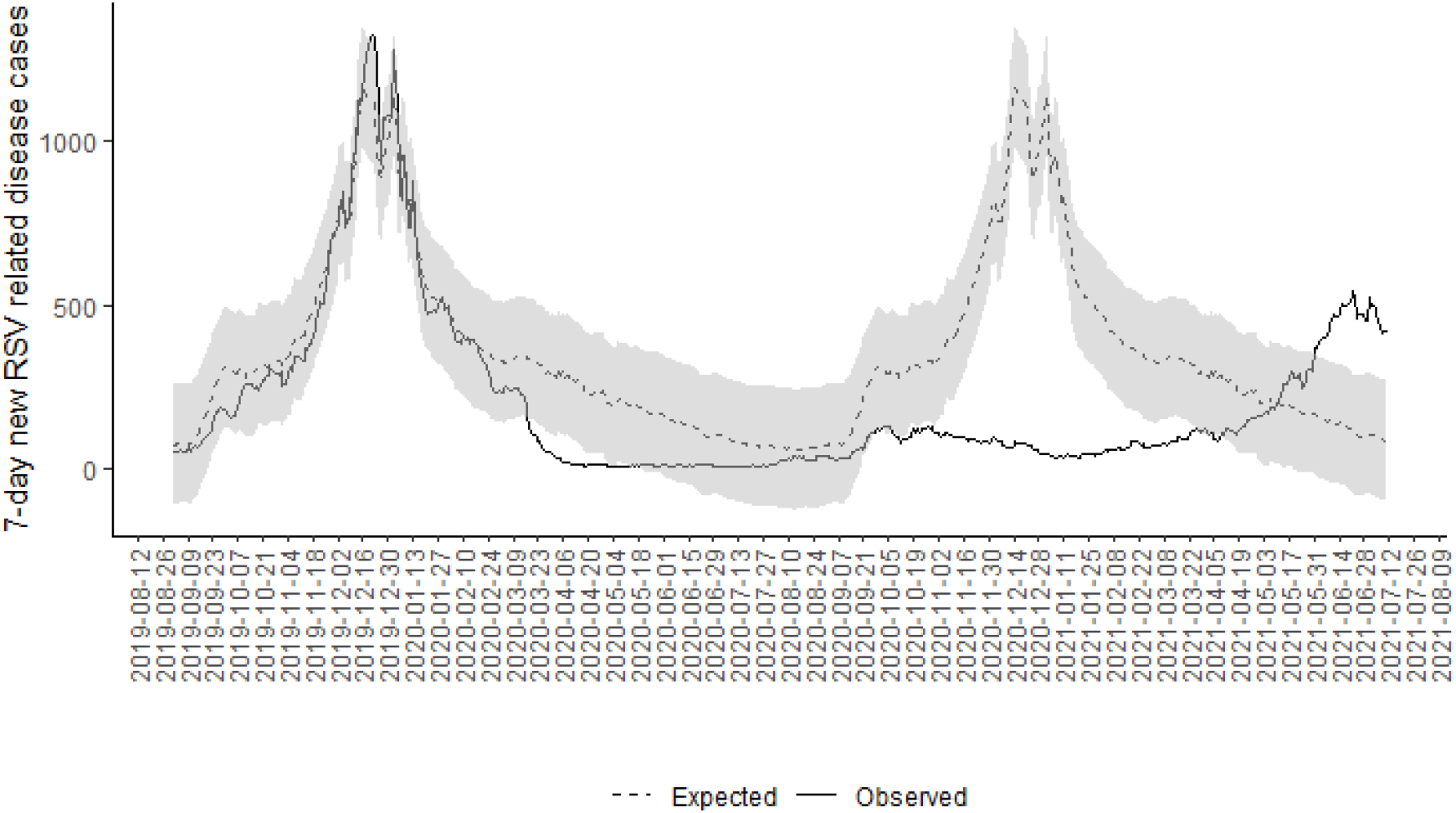
**Observed and expected (with 95% CI) new RSV-related disease cases in a 7-day period from September 2019 to July 2021 in Catalonia in children under 5 years old.**

Finally, we performed the same analysis with RSV laboratory-confirmation data. We observed that laboratory-confirmations in the 2019-2020 season were as expected. During the 2020-2021 winter, however, only one sporadic laboratory-confirmation was observed until week 8 2021 and 608 RSV were thereafter confirmed between weeks 9 and 26, showing the same pattern as the EHR data (Figure 2). Regarding testing, 2020-2021 season nearly doubles that of the 8 previous seasons average and the percentage of samples with positive results for RSV were 11% in the current season versus an average of 15.8% for the previous seasons (Supplementary Table S2).

**Figure 2.**
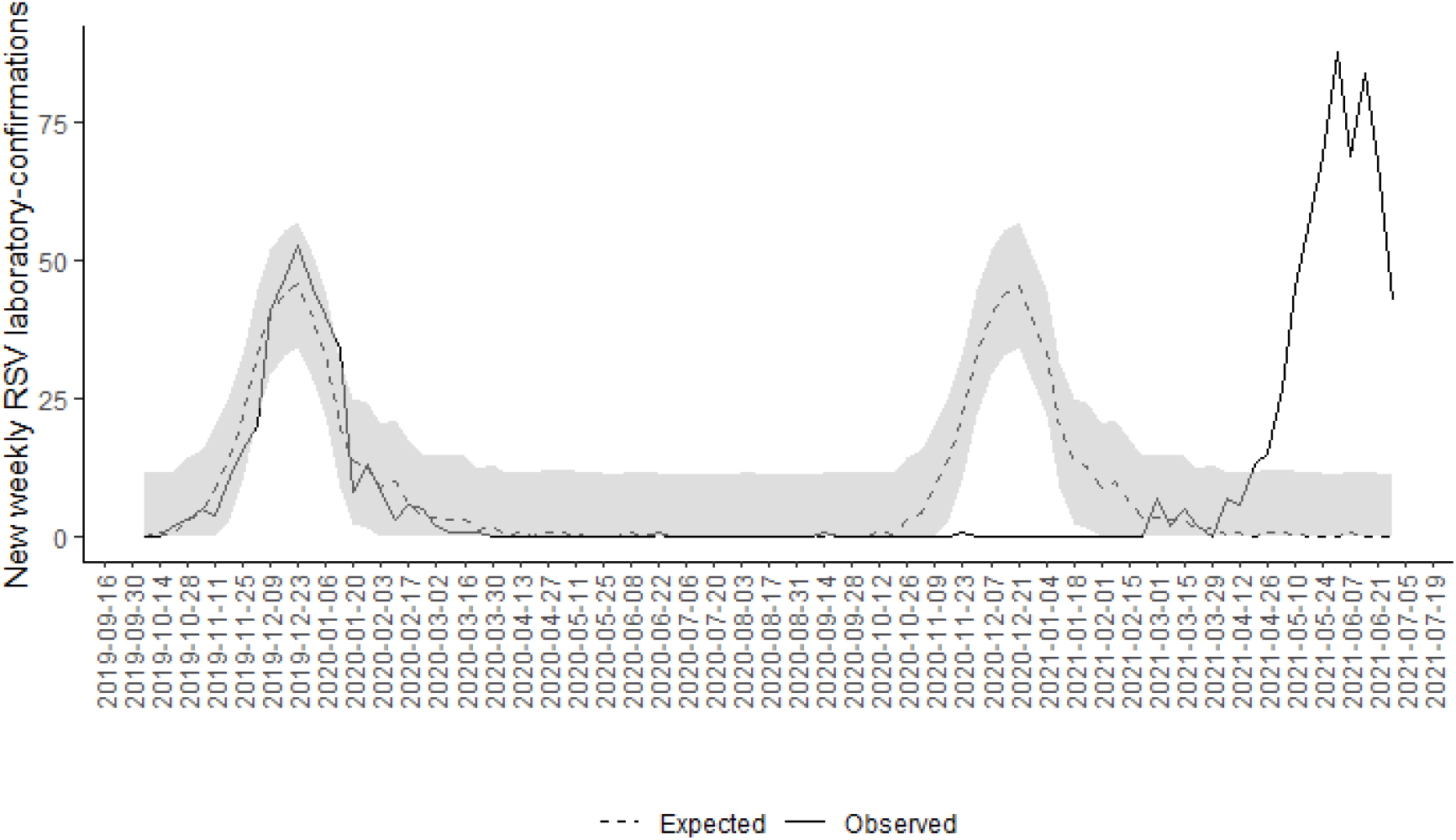
**Observed and expected (with 95% CI) new weekly RSV laboratory-confirmations from October 2019 to July 2021 in Catalonia.**

## Discussion

We observed an unusual trend of RSV and bronchiolitis disease during the 2020-2021 season. RSV circulation was nearly absent during the winter and a resurgence was reported in late spring, similar to ECDC data from Iceland and France in the Northern hemisphere, and consistent with previous data from Australia, in the Southern hemisphere [3, 4].

Researchers in Australia suggested that both RSV reduction and posterior rise of cases could be related to non-pharmaceutical control measures implemented to contain COVID-19 [3,4]. In Spain, a second state of alarm was in place from 25 October 2020 to 9 May 2021, which allowed several restrictions in the community, including different levels of perimeter lockdowns and curfew from 10 pm to 6 am. However, schools and pre-schools remained open from September to June 22 (last day of scholar year in Catalonia) with some mitigation measures including bubble groups, hygiene measures, daily symptoms screening and mandatory face-mask for children older than 6 years [8].

Although these measures to control SARS-CoV-2 transmission could have an impact on other respiratory viruses, it is less clear that they explain the resurgence during the spring, as RSV was already on its rise before lifting the main measures. It is possible that the lower circulation during winter led to less immunization in younger children and thus increased their individual susceptibility and the number of susceptibles over time [9]. Other causes should also be considered. For instance, it has been hypothesized that changes in patients’ health-seeking behaviour in the context of COVID-19 may have contributed to changes in RSV activity [10]. Finally, it is also possible that viral interference between SARS-CoV-2 and other respiratory viruses plays a role in this unusual pattern of RSV [11]. More studies are needed to accurately test the different hypotheses and to analyze if SARS-CoV-2 will affect and displace the peak of other seasonal viruses. Despite this peak in late spring and early summer, it is unpredictable whether the next RSV or bronchiolitis epidemic will occur during the fall, when usual.

Although the laboratory-confirmation data pattern shows similar timing to EHR data, the resurgence peak was higher than in previous winter peaks. In this sense, the hospital laboratory is testing many more samples than in previous seasons due to the need for SARS-CoV-2 screening, even though these cases do not present clinical criteria for the hospital admission. Since RSV and other respiratory viruses are tested systematically in respiratory samples collected from acute respiratory infection cases, RSV might be overdiagnosed in the last months. This change allows for the recent increase in RSV testing and absolute detections while decreasing the percentage of positive results for RSV.

Among the limitations of our study, we should address the use of clinical diagnoses from primary care EHR for surveillance, as some studies suggested that clinical codes could underestimate the number of real RSV infections [6]. However, our results were consistent with the virus laboratory-confirmations data from one of the biggest paediatric hospitals in Spain. Continued collaborative surveillance of RSV and related disease in primary care and hospital settings could help to detect early changes in trends and to prompt public health data-driven decision-making, becoming a warning system of unexpected patterns in the coming months, as well as to monitor mild and severe RSV-related disease.

## Data Availability

EHR data and analytical code are provided at https://github.com/ErmengolComa/RSV_.git . Laboratory-confirmation data are available only under reasonable request.

https://github.com/ErmengolComa/RSV_.git

## Additional sections

### Ethics approval

This study was done in accordance with existing statutory and ethical approvals from the Clinical Research Ethics Committee of the IDIAPJGol (project code: 20/172-PCV) and the Vall d’Hebron Hospital Campus Clinical Research Ethics Committee (PR(AMI)556/2020).

### Funding

AA and JV’s work was partially supported by national Plan of R + D + I 2008–2011 and by Carlos III Health Institute, Subdirectorate-General of networks and cooperative research Centers, Spanish Ministry of Economy and Competitiveness, Spanish Network for Research in Infectious Diseases [REIPI RD16/0016/0003]; By Health Research Fund, Spanish Ministry of Economy and Competitiveness [grants FIS PI14/01838 and PI18/00685]; and European Regional Development Fund (ERDF).

### Data availability statement

EHR data and analytical code are provided at https://github.com/ErmengolComa/RSV_.git. Laboratory-confirmation data are available only under reasonable request.

## Acknowledgments

The authors of this paper would like to thank all health care professionals in Catalonia for their work and resilience providing care to the Catalan population during these difficult times. We also would like to acknowledge the efforts of all members of the SISAP team during the last months.

## Authors’ contributions

All authors contributed to the design of the study, the interpretation of the results, and reviewed the manuscript. EC,LM and NM had access to the EHR data and acted as guarantors. AA had access to the laboratory-confirmation data. NM performed the statistical analysis. EC, AA, LM and JV wrote the first draft of the manuscript. All authors critically revised the manuscript. The corresponding author attests that all listed authors meet authorship criteria and that no others meeting the criteria have been omitted.

