## Supplementary material for "Unusual trend of respiratory syncytial virus burden of disease observed in primary care diagnosis of children under 5 years old in Catalonia during the COVID-19 pandemic"

**Supplementary Figure S1. Trends of RSV-related disease diagnosis from September 2009 in children under 5 years old.**

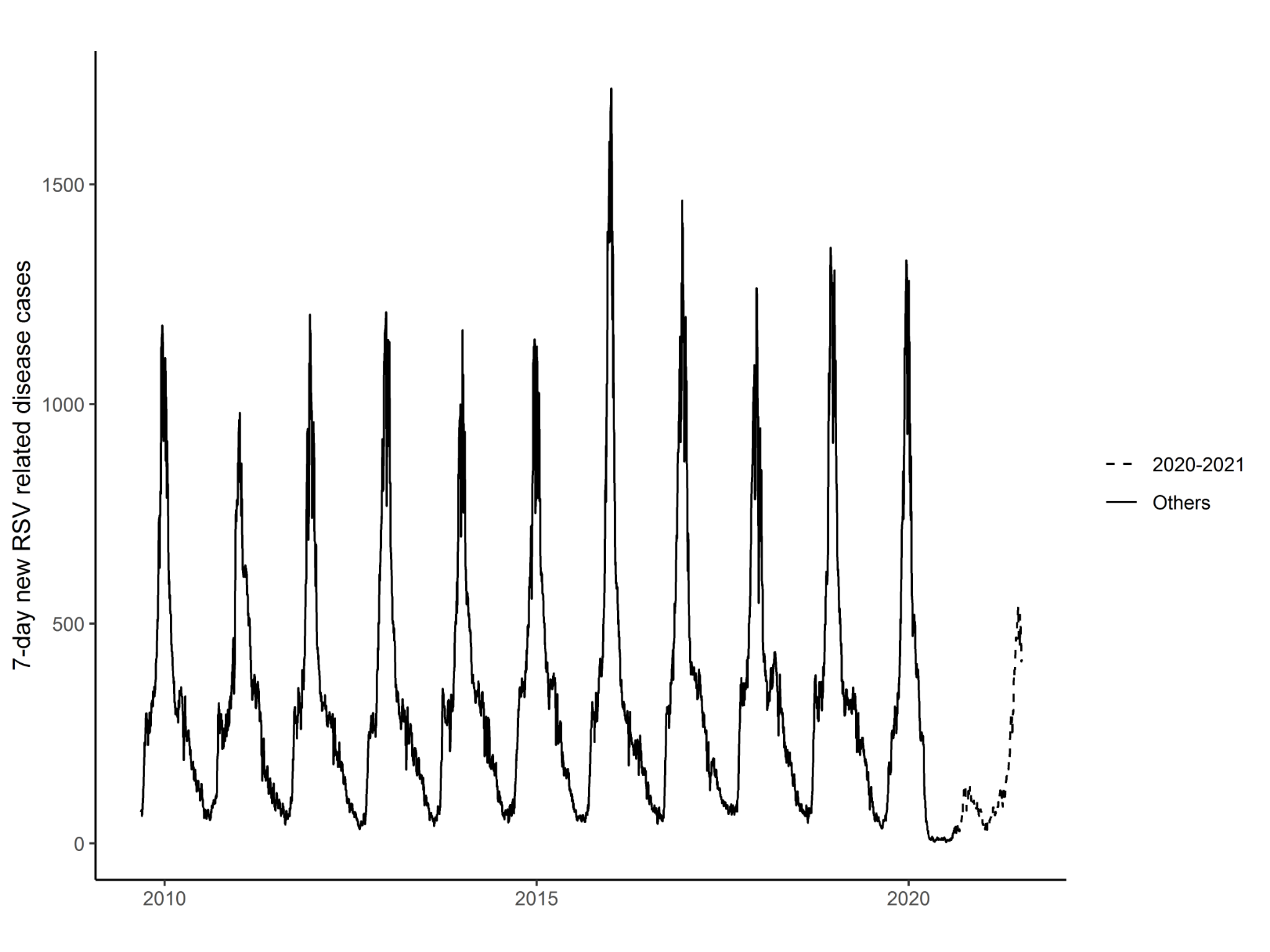

**Supplementary Table S1. Number of RSV diagnoses by age group from 2009-2010 season to 2020-2021 season (until 4th July)**

| **Age group** | **2009 - 2010** | **2010 - 2011** | **2011 - 2012** | **2012 - 2013** | **2013 - 2014** | **2014 - 2015** | **2015 - 2016** | **2016 - 2017** | **2017 - 2018** | **2018 - 2019** | **2019 - 2020** | **2020 - 2021** |
| --- | --- | --- | --- | --- | --- | --- | --- | --- | --- | --- | --- | --- |
| **<2** | 16,004  (93.8%) | 15,110  (93.9%) | 14,952  (94.6%) | 16,056  (93.8%) | 15,689  (93.3%) | 16,738  (93.7%) | 18,546  (93.9%) | 17,630  (94.9%) | 17,179  (94.2%) | 17,556  (93.8%) | 12,901  (95.3%) | 6,022  (90.1%) |
| **Between 2-4** | 1,059  (6.2%) | 983  (6.1%) | 847  (5.4%) | 1,067  (6.2%) | 1,120  (6.7%) | 1,121  (6.3%) | 1,214  (6.1%) | 957  (5.2%) | 1,054  (5.8%) | 1,169  (6.2%) | 639  (4.7%) | 664  (9.9%) |
| **Total** | **17,063** | **16,093** | **15,799** | **17,123** | **16,809** | **17,859** | **19,760** | **18,587** | **18,233** | **18,725** | **13,540** | **6,686** |

**Supplementary Table S2. Number of samples, RSV laboratory-confirmations and percentage of positive samples by season**

| **Season** | **Samples** | **RSV+** | **% of positives** |
| --- | --- | --- | --- |
| 2012-2013 | 1,989 | 376 | 18.9% |
| 2013-2014 | 1,913 | 294 | 15.4% |
| 2014-2015 | 2,516 | 361 | 14.3% |
| 2015-2016 | 2,796 | 427 | 15.3% |
| 2016-2017 | 2,567 | 437 | 17.0% |
| 2017-2018 | 2,624 | 447 | 17.0% |
| 2018-2019 | 2,665 | 422 | 15.8% |
| 2019-2020 | 2,790 | 371 | 13.3% |
| **2012-2020** | **19,860** | **3,135** | **15.8%** |
| 2020-2021 | 5,479 | 609 | 11.1% |
